# Nasal septum deviation correlates with asymmetry in Parkinson’s disease

**DOI:** 10.1101/2025.02.27.25322941

**Authors:** Jing Wang, Fengtao Liu, Jingjie Ge, Yimin Sun, Yilin Tang, Xiaoniu Liang, Yixin Zhao, Wenbo Yu, Jianjun Wu, Chensen Lin, Yuanye Zhou, Yichi Zhang, Jianning Zhang, Ping Wu, Jiaying Lu, Zizhao Ju, Yuhua Zhu, Huamei Lin, Yunhao Yang, Qian Xu, Wei Zheng, Jianfeng Luo, Yihui Guan, Honglei Chen, Mei Tian, Chuantao Zuo, Jian Wang

## Abstract

**Background:** Asymmetry in motor dysfunction and associated dopaminergic deficit is a common characteristic of Parkinson’s disease (PD), yet potential explanations remain mysterious. Hereby, we assessed whether asymmetry in the nasal cavity is related to dopaminergic dysfunction asymmetry in PD patients.

**Methods:** This cross-sectional, multi-center observational study included 761 PD patients from three cohorts. First, we analyzed data from the Huashan Parkinsonian PET Imaging Database (March 2011 to February 2020), which served as the primary cohort (n=333). Second, we collected de novo data from all PD inpatients in the Hongqiao Campus of Huashan Hospital as internal validation cohort (May 2023 to July 2024, n=77). Finally, we used data from the Parkinson’s Progression Markers Initiative as an external validation cohort (n=351). All cohorts included imaging data of structural MRI or CT, as well as dopaminergic neuroimaging using ^11^C−CFT, ^18^F−FP−CIT, or ^18^F−DTBZ on PET or ^123^I−DaTscan on SPECT. Nasal cavity asymmetry was assessed by visually inspecting the position of nasal septum deviation in structural MRI or CT to determine the dominant side. Both qualitative and quantitative analyses were performed to evaluate the correlations between nasal cavity asymmetry and dopaminergic deficit asymmetry.

**Results:** In the primary cohort, 70.2% of patients exhibited consistency between the dominant side of the nasal cavity and the side with a more severe dopaminergic deficit in striatum (φ=0.40, p<0.001). The striatal−specific binding ratios of dopamine uptake were significantly lower on the side ipsilateral to the dominant nasal cavity, and a significantly inverse correlation was found between the asymmetry index of nasal cavity surface area and that of the dopaminergic deficit (r=−0.31, p<0.001). Similar patterns were observed in internal (78.6%, φ=0.57, p<0.001) and external validation cohorts (73.1%, φ=0.45, p<0.001). A stronger correlation was found in sporadic PD (φ=0.67, p<0.001) compared to genetic PD patients (φ=0.31, p=0.3).

**Conclusions:** We made a novel and robust observation that the nasal cavity asymmetry is correlated with asymmetry in striatal dopaminergic deficiency in PD patients. The finding may have significant implications for both the etiological and clinical research of PD, supporting the nasal pathway as a potential route for both environmental pathogens and PD treatment.

**Trial Registration** Chinese Clinical Trial Registry, ChiCTR2400094198, and https://www.chictr.org.cn/bin/project/edit?pid=251807

## Introduction

Parkinson’s disease (PD), a complex neurodegenerative disease with a wide spectrum of clinical manifestations, is primarily characterized by resting tremor, bradykinesia, and rigidity. A hallmark feature of these symptoms is their prominent asymmetry, which is often considered an important diagnostic clue in the Movement Disorder Society (MDS) diagnostic criteria for PD.^1^ This asymmetry in clinical presentation is believed to reflect an underlying asymmetry in pathological neuronal loss, particularly within dopaminergic pathways. However, the reasons underlying for this symptomatic asymmetry remain elusive.^2,3^ Deciphering this phenomenon could provide critical insights into the etiology of PD and inform the development of novel treatment strategies.^4^

Environmental exposures, either alone or interacting with genetics, may contribute to the development and progression of PD.^5,6^ Notably, the nasal olfactory pathway has been implicated as a route of exposure, primary because it is one of the body’s main interfaces with the environment.^7^ Hyposmia, or the loss of the sense of smell, is one of the most notable prodromal symptoms of PD, further supporting this route. Airborne pollutants have been associated with an increased PD risk, and experimental studies have shown that long-term nasal intranasal instillation of lipopolysaccharides (LPS) can lead to nigro−striatal dopaminergic degeneration.^8^ While preliminary, these evidence highlight the importance of studying the nasal pathway as a potential route involved in the environmental etiology of PD.

We further speculate that structural asymmetry of the nasal cavity may influence environmental exposures to neurotoxicants, potentially contributing to asymmetry in dopaminergic neuronal degeneration and symptom presentation. A deviated nasal septum, which is observed in approximately 90% of adults,^9^ is typically considered a benign developmental anatomical variation but can lead to asymmetric exposure to environmental pathogens due to differential airflow between nasal cavities. This deviation results in increased airflow and potential pathogen exposure in the dominant, or larger, side of the nasal cavity. Therefore, this study hypothesizes that increased neurotoxicant exposure in the dominant nasal cavity may, over time, cause greater damages to the ipsilateral nigrostriatal system through the nose−to−brain route, ultimately contributing to PD−related pathological and clinical asymmetry.

## Methods

### Study design and participants

This cross−sectional, multi−center study included PD patients from three independent cohorts. In the primary cohort, the correlation between nasal cavity asymmetry and asymmetry in dopaminergic dysfunction in the putamen was accessed using both qualitative and quantitative methods. Data were obtained from the Huashan Parkinsonian PET Imaging Database,^10^ which recruited PD patients from March 2011to February 2020 at the Gamma Campus of Huashan Hospital in Shanghai, China. To validate the findings from the primary cohort and reduce potential selection bias, an internal validation cohort consisting of consecutively hospitalized PD patients at the Hongqiao Campus of Huashan Hospital from May 2023 to July 2024 was used.

An external validation cohort from the Parkinson’s Progression Markers Initiative (PPMI) was used to further test the robustness and generalizability of findings. This cohort included PD patients who underwent ^123^I−DaTscan SPECT and structural MRI or CT scans between January 2011 and December 2023. PPMI data are publicly available at https://www.ppmi-info.org with clearly stated inclusion and exclusion criteria. This cohort was selected to ensure an independent and externally valid test of the primary and internal cohort findings, particularly given the diverse nature of data sources and imaging modalities.

This study was approved by the Institutional Review Boards of Huashan Hospital (KY2024−1295). All study participants provided written consent. This study followed the Strengthening the Reporting of Observational Studies in Epidemiology (STROBE) reporting guideline.

### Procedure

Participants from the primary and internal validation cohorts were evaluated by movement disorder specialists, using the MDS−Unified Parkinson’s Disease Rating Scale (MDS−UPDRS) parts III (motor examination; recorded in the off−state for treated participants) and the Hoehn−Yahr stage. Patients enrolled before 2015 were diagnosed using the United Kingdom Brain Bank Criteria, and those diagnosed afterwards met the MDS PD criteria. All participants were at least 30 years old at the time of diagnosis.

Dopaminergic dysfunction (DAT or VMAT2 binding) in the striatum was assessed using ^11^C−CFT or ^18^F−DTBZ PET/CT scans in the primary cohort and ^18^F−FP CIT PET/CT scans in the internal validation cohort. All participants exhibited abnormal dopamine deficits. Detailed imaging acquisition protocols and neuroimaging processing methods are provided in appendix (pp 2).

Putamen−specific binding ratios (SBRs) were calculated as putamen/occipital−1, focusing on the putamen as nigrostriatal degeneration affects it earlier than the caudate nucleus. An asymmetry index (AI) was derived to quantify the degree of asymmetry in putaminal dopaminergic deficit: 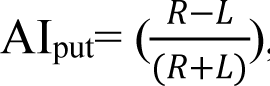, where R and L represent right and left putamen SBRs. Thresholds for significant asymmetry were AI_put_≤–0.04 or AI_put_≥0.04 for the primary and internal validation cohorts, based on 176 healthy controls (as described in appendix pp 3), and AI_put_≤–0.08 or AI_put_≥0.08 for the external validation cohort as reported by Knudsen et al. (as described in appendix pp 3). Different cutoffs were used in these cohorts may due to differences in imaging tracers.

Two accredited clinicians independently examed nasal CT or MRI in a blinded manner to determine the dominant side of nasal cavity. In case of disagreement, a third expert participated in the review and the final decision was made by consensus. Nasal septum deviation was qualitatively categorized into three groups: straight septum (no deviation), S−shaped deviation (i.e., bending in both directions), and unilateral deviation (i.e., deviating to either the left or right side). Visual reading methods and results are provided in appendix (pp 4–5).

In addition to qualitative measures of nasal cavity asymmetry, we also quantitatively measured the surface area and volume for both sides of the nasal cavity based on CT images.^11^ Briefly, we first identified nasal cavity segmentation with manual annotation and a 3D convolutional neural network, followed by visual inspection of the segmented data. In case of segmentation errors, manual adjustments were made. Measurements were then extracted using 3D Slicer 5.6.2 (Radiomics; Asan Medical Center Nuclear Medicine Image Quantification Toolkit of Excellence), a software available at https://www.slicer.org/. Detailed methodology is outlined in appendix (pp 6–7). The AI calculation formula was used to quantify the degree of asymmetry in the nasal cavity, where R and L represent the right and left nasal cavity phenotypes (surface area and volume). Finally, computational fluid dynamics (CFD) was used to simulate the patient’s nasal exposure path to fine particulate matter less than 2.5μm in diameter and predict major deposition locations (as described in appendix pp 9).

### Outcomes

The primary outcome of this study was to assess the correlation between the dominant nasal cavity side, due to nasal septum deviation, and the striatal side with more pronounced dopaminergic deficits in PD patients. Secondary outcomes included comparing dopamine levels in the striatum between the dominant nasal cavity side and the contralateral side, if the dominant nasal side corresponds to more severe dopaminergic deficits. Additionally, the study analyzed the Pearson correlation between the AI of nasal cavity and the AI of striatal damage to evaluate the degree of association between the two in PD patients.

### Statistical analysis

Descriptive analysis included means ± standard deviations for continuous variables and proportions for categorical variables. The Phi coefficient (φ) was used to assess the correlation between the dominant side of the nasal cavity and the side with more putamen dopaminergic deficits (as described in appendix pp 10). The Wilcoxon matched−pairs signed−rank test, a non−parametric method, was used to analyze SBRs differences between the dominant and contralateral sides. Pearson correlation coefficients between the AI of putaminal dopaminergic deficits and those of nasal cavity surface area and volume were calculated, with all correlations were Fisher z–transformed.

All statistical analyses were performed using SPSS software (version 27.0; IBM, Armonk, NY, USA), Origin (Version 2024b; Origin Lab Corporation, Northampton, MA, USA), and R software (v.4.2.2) utilizing the ggplot2 package (v.3.4.2) for graph creation. Two−tailed p values of less than 0.05 were considered statistically significant.

## Results

### Study Population

The study included a total of 761 participants: 333 from the primary cohort, 77 from the internal validation cohort, and 351 from the external validation PPMI cohort (figure 1). Participant demographics are summarized in table 1. eFigure 7 (appendix pp 11) shows the distribution of different patterns of nasal septum deviation in PD patients with pronounced striatal dopaminergic asymmetry across three cohorts, illustrating their overall relationship with dopaminergic asymmetry.

**Table 1:** Demographic and clinical characteristics from primary cohort, internal validation cohort, and external validation cohort. MDS−UPDRS III=MDS Unified Parkinson Disease Rating Scale, Part III. NA=No applicable

|  | Primary cohort (n = 333) | Internal validation cohort (n = 77) | External validation (PPMI) cohort (n = 351) |
| --- | --- | --- | --- |
| Age, median (IQR), y |  |  |  |
| Sex, No. (%) |  |  |  |
| Male | 206 (62) | 42 (55) | 203 (58) |
| Female | 127 (38) | 35 (45) | 148 (42) |
| Disease duration, median (IQR), y | 2 (1 to 3.8) | 2.6 (1.1 to 5.2) | NA |
| MDS–UPDRS III total score, median (IQR) | 19 (13 to 27) | 35 (20 to 45) | 21 (16 to 29) |
| Hoehn–Yahr stage, median (IQR) | 2 (1 to 2) | 2 (2 to 3) | 2 (1 to 2) |
| No. of Dopaminergic PET imaging |  |  |  |
| <sup>11</sup>C–CFT | 239 | 0 | 0 |
| <sup>18</sup>F–FP CIT | 0 | 77 | 0 |
| <sup>18</sup>F–DTBZ | 94 | 0 | 0 |
| <sup>123</sup>I–DaTscan | 0 | 0 | 351 |
| No. of Structure imaging |  |  |  |
| CT | 333 | 73 | 31 |
| MRI | 0 | 4 | 320 |
| No. of Genetic test | NA | NA | 153 |
| None gene identified | NA | NA | 115 |
| LRRK2 variant | NA | NA | 17 |
| GBA variant | NA | NA | 18 |
| PRKN variant | NA | NA | 2 |
| LRRK2 & GBA variant | NA | NA | 1 |

**Figure 1:**
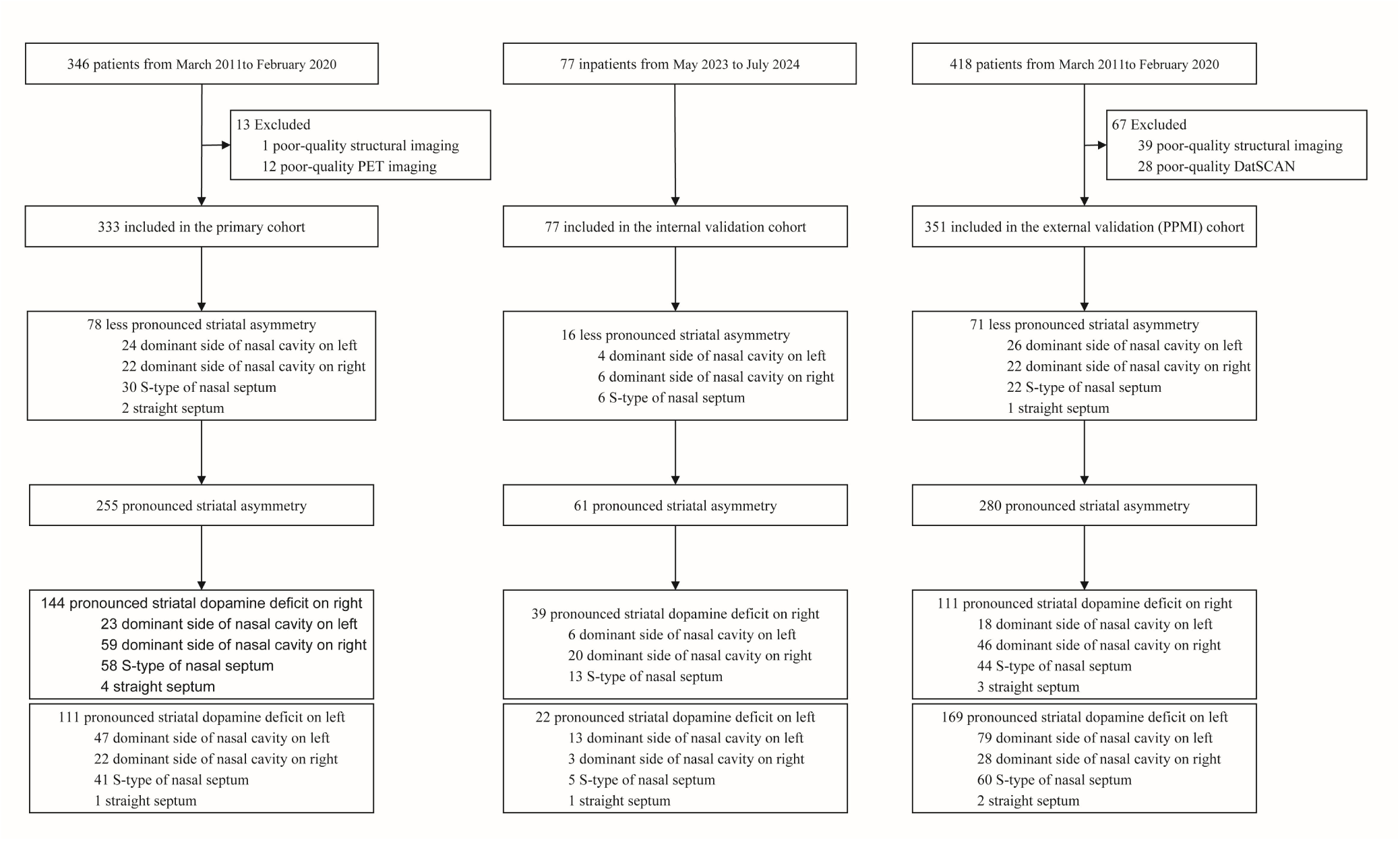
Flow of study participants

### Laterality Relationship Between Nasal Cavity Dominance and Striatal Dopaminergic Deficit

In the primary cohort, 255 patients demonstrated pronounced striatal asymmetry of dopaminergic deficits, among whom 151 exhibited unilateral nasal septum deviation. Of these 151 patients, 106 (70.2%) showed a pattern consistent with our hypothesis: a more severe striatal dopaminergic deficit corresponded to the dominant side of the nasal cavity (table 2). Qualitative analysis revealed a significant positive correlation between the dominant nasal cavity side and the side with more pronounced striatal dopaminergic deficit (φ=0.40, p<0.001). This correlation was stronger in patients with early−stage PD (Hoehn−Yahr stage=1, φ=0.48, p<0.001; eTable 1, appendix pp 12).

**Table 2:**
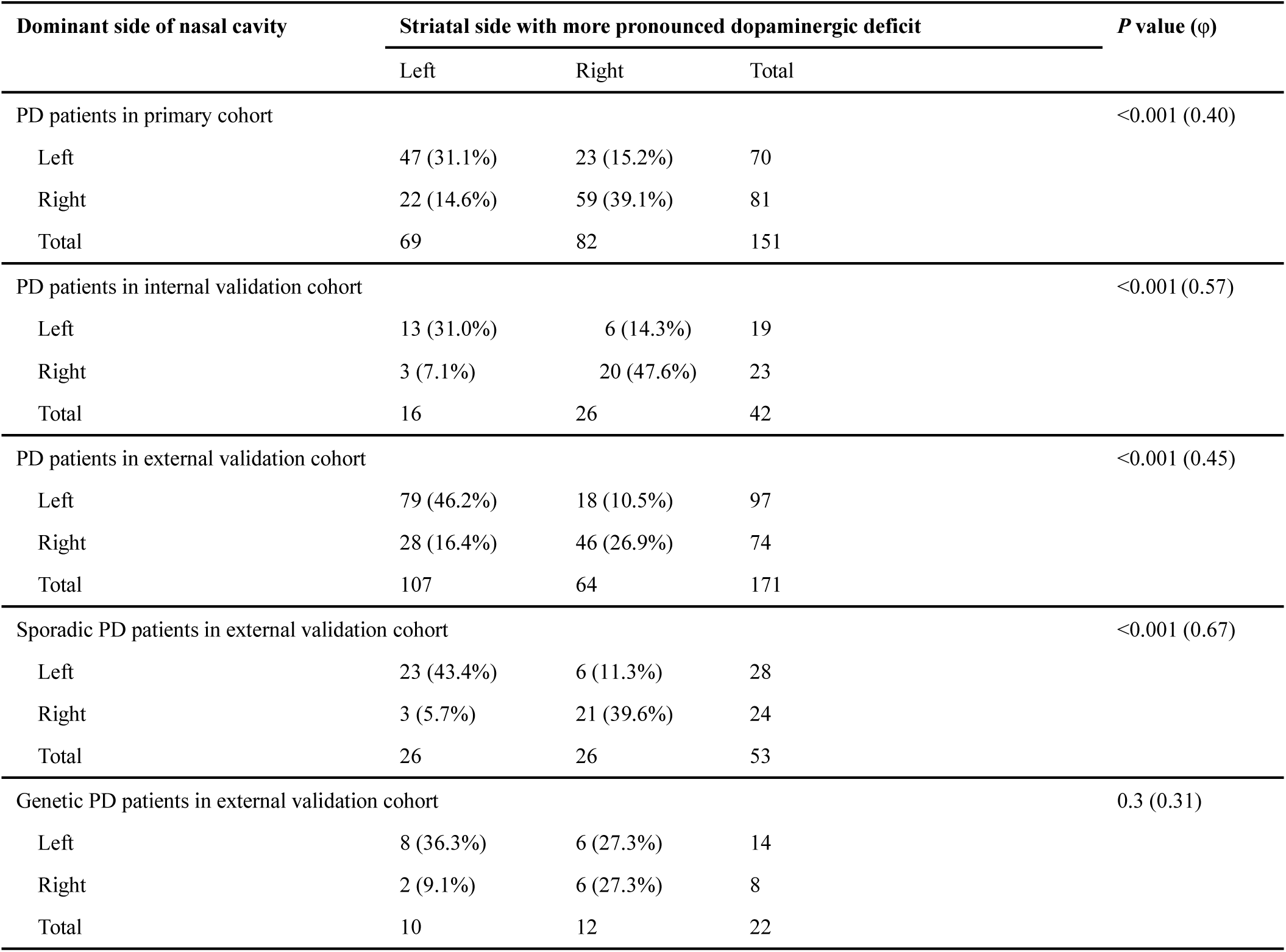
The correlation between the dominant side of nasal cavity and the striatal side of more pronounced dopaminergic deficit across cohorts. PD=Parkinson’s disease

In the internal validation cohort, 61 patients displayed pronounced striatal asymmetry, and 42 had unilateral nasal septum deviation (19 left−dominant, 23 right−dominant). Among these 42 patients, 33 (78.6%) exhibited patterns consistent with our hypothesis, showing more severe dopaminergic deficits on the dominant side of nasal cavity (φ=0.57, p<0.001, table 2).

In the external validation cohort, 280 of 351 PD patients demonstrated pronounced striatal asymmetry, with 171 showing unilateral nasal septum deviation (97 left−dominant, 74 right−dominant). Among these, 125 (73.1%) exhibited asymmetry patterns consistent with our hypothesis (table 2). Further analysis confirmed a significant positive correlation between the dominant nasal cavity side and the side with more severe striatal dopaminergic deficit (φ=0.45, p<0.001). Of the 351 PD patients, 153 underwent genetic testing (table 1),^12^ including 115 classified as sporadic PD and 38 as genetic PD. Subgroup analysis showed a stronger correlation in sporadic PD (φ=0.67, p<0.001) compared to genetic PD (φ=0.31, p=0.3; table 2).

### Difference in Putamenal Dopaminergic Deficit Between Nasal Cavity Dominant and Contralateral Sides

In the primary cohort, putamenal SBRs were significantly lower on the dominant side of the nasal cavity compared to the contralateral side (Wilcoxon matched−pairs signed−rank test, p<0.001; figure 2A). This finding was replicated in the internal validation cohort (p<0.01; figure 2B) and further validated in the external validation cohort (p<0.001; figure 2C).

**Figure 2:**
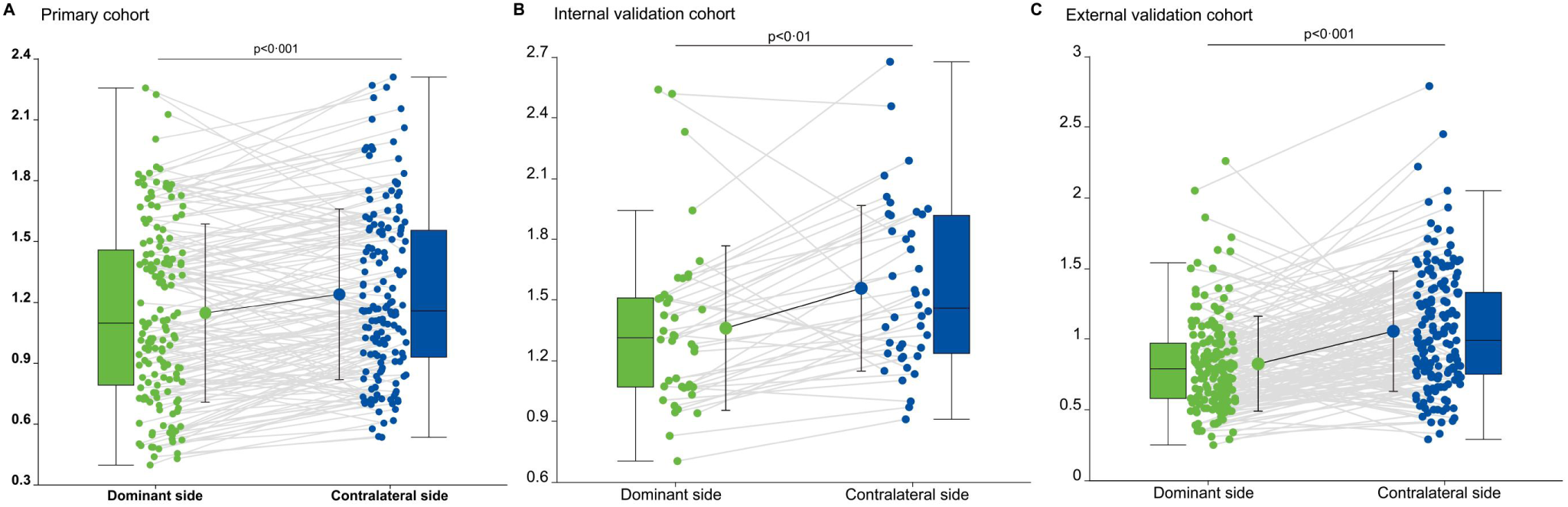
The putamen specific binding ratios on the dominant side and the contralateral side of nasal cavity. Striatal specific binding ratios on the dominant side of nasal cavity were significantly lower than that on the contralateral side in primary cohort (A), internal validation (B), and external validation cohort (C) as shown by Wilcoxon matched−pairs signed−rank tests. SBRs=specific binding ratios

### Correlation Between Nasal Cavity Laterality Index and Striatal Dopaminergic Uptake

A statistically significant inverse correlation was further identified between AI_put_ of dopamine uptake and the LI of nasal cavity phenotypes in the primary cohort (Pearson’s correlation coefficients: surface area with AI_put_: r=−0.31 [95%CI, −0.44 to −0.16], p<0.001, figure 3; volume with AI_put_: r=−0.24 [95%CI, −0.38 to −0.09], p<0.01, eFigure 8 in appendix pp 13, respectively).

**Figure 3:**
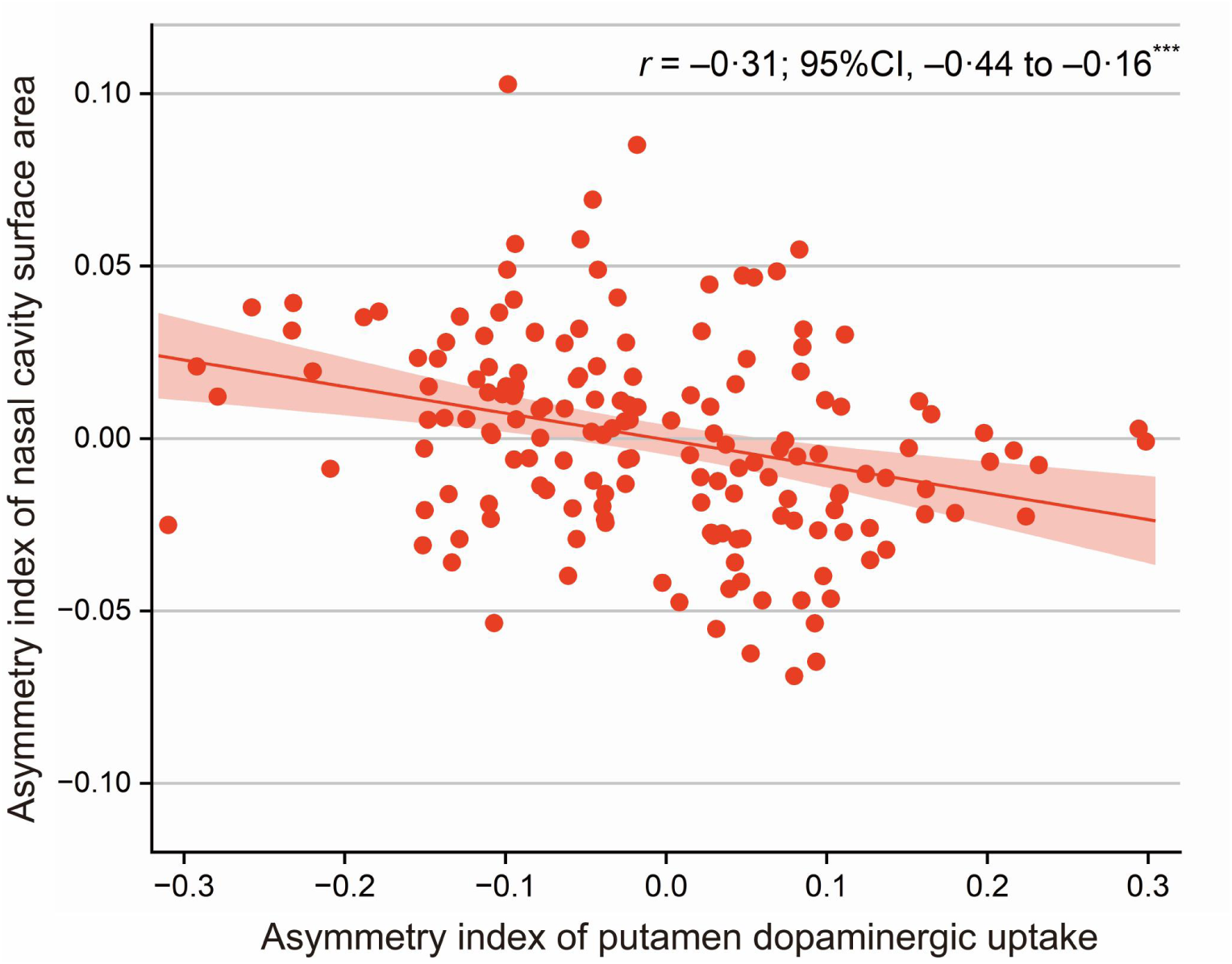
Association between the laterality index of nasal cavity surface area and laterality Index of putamen dopaminergic uptake in the primary cohort *** p<0.001

### Nasal Computational Fluid Dynamics Simulation

Figure 4 visualizes the patterns of putaminal dopaminergic loss and the dominant side of nasal cavity in PET/CT scans of PD patients with deviated nasal septum. The airflow results for the left and right nasal cavities are shown in figure 5, demonstrating that the septal deviation creates distinct airflow patterns in each cavity. Streamline analysis reveals that the upper portion of the left nasal cavity primarily resides in a slow recirculation zone with minimal airflow passing through. In contrast, a substantial volume of external air flows continuously through the upper part (olfactory region) of the right nasal cavity. This airflow pattern significantly affects particle deposition, with results showing markedly less particle deposition in the upper part of the left nasal cavity. Additionally, this video (https://drive.google.com/file/d/1TOyC6seXknQcAzqr-vhy2t-sblZBgsSe/view?usp=sharing, appendix media) simulates a patient’s nasal exposure to particles, highlighting potential deposition sites.

**Figure 4:**
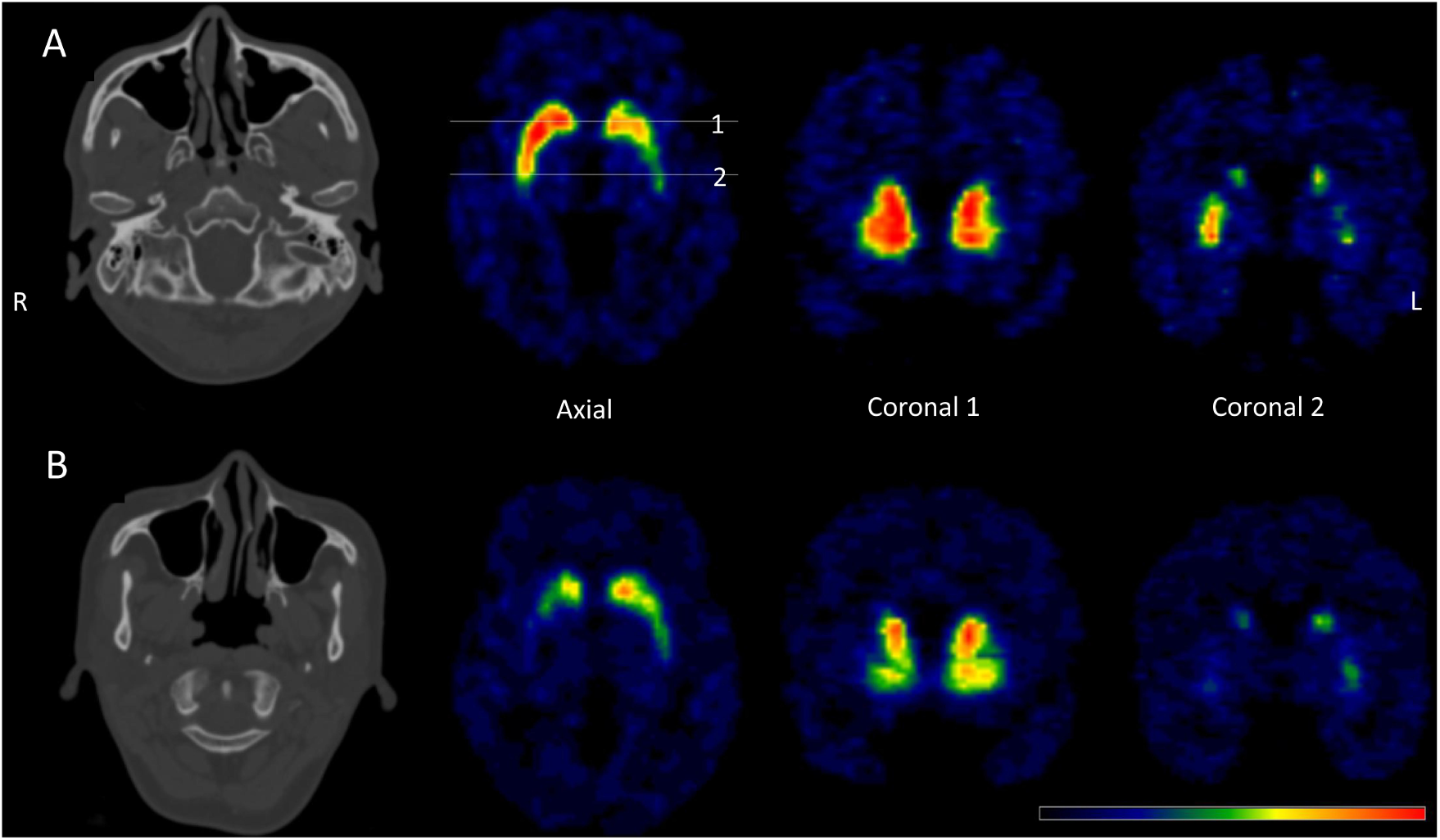
The patterns of striatal dopamine loss based on ^11^C−CFT PET/CT in two Parkinson’s disease patients with deviated nasal septum. (A) One patient with the dominant side of nasal cavity on the left due to a deviated nasal septum showed more pronounced dopaminergic loss in the left putamen (Patient A). (B) One patient with the dominant side on the right due to a deviated nasal septum showed more pronounced dopaminergic loss in the right putamen (Patient B).

**Figure 5:**
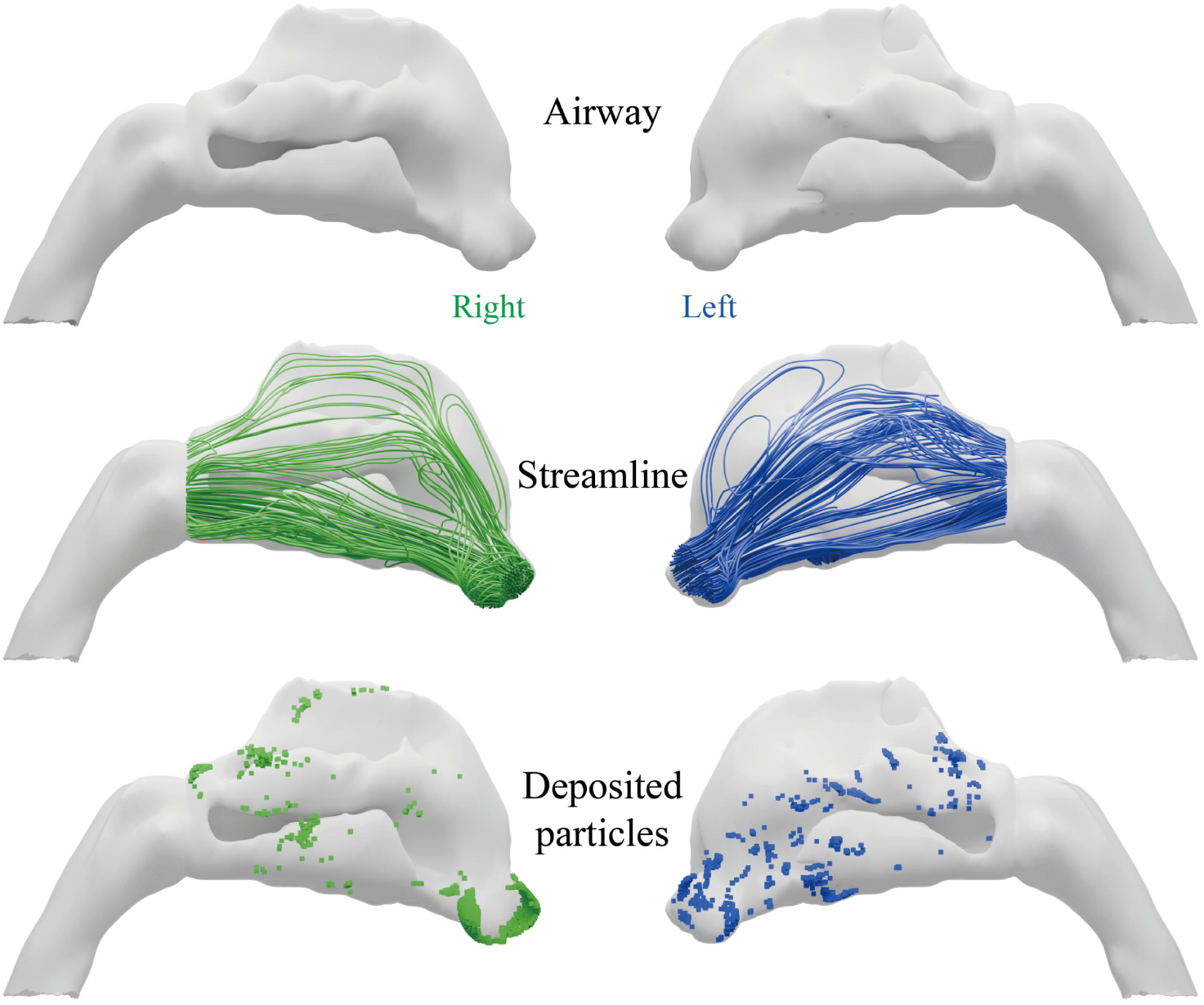
Computational fluid dynamics simulation of the streamline and deposited particles in patient’s nasal cavity. This illustration shows Patient B, who has a deviated nasal septum with a dominant side on the right. The airflow results for the left and right nasal cavities are represented in blue and green, respectively. Streamline analysis reveals that a substantial volume of external air flows continuously through the upper part (olfactory region) of the right nasal cavity. This airflow pattern significantly affects particle deposition, with results showing markedly less particle deposition in the upper part of the left nasal cavity (blue squares) compared to the right (green squares).

## Discussion

To our knowledge, this is the first study linking nasal cavity asymmetry to dopaminergic deficits asymmetry in PD patients. While preliminary, this finding is remarkably robust across three independent cohorts with drastically different study settings, supporting its validity and generalizability. Specifically, in all three cohorts, most patients displayed more pronounced dopaminergic deficits in the putamen on the same side as the dominant nasal cavity. These novel observations, combined with the increasingly recognized nose−to−brain pathway in PD pathogenesis, could provide crucial insights into the role of airborne environmental exposures in PD and inform the development of novel treatments targeting the nasal route.

The symmetric clinical onset and dopaminergic deficit in PD have been extensively studied, yet the underlying causes remain elusive. One hypothesis suggests hemispheric differences in brain region volume, shape, or the susceptibility of dopaminergic neurons in the substantia nigra, leading to accelerated degeneration on one side.^13^ However, these hypotheses lack strong pathological evidence.^2^ It has also been proposed that hemispheric differences in DNA methylation patterns may influence neuronal degeneration in PD,^14^ but supporting evidence remains limited. With empirical data from three independent patient cohorts, our findings raise the intriguing possibility that nasal cavity asymmetry may be a contributing factor. The larger side of the nasal cavity may be more exposed to environmental neurotoxicants, facilitating their entry into the brain through the olfactory nerve pathway and potentially initiating lateralized dopaminergic degeneration in the substantia nigra. This novel hypothesis, though provocative, is well supported by the increasing recognition of the nose-to-brain pathway’s role in PD pathogenesis.

Environmental neurotoxicants, such as air pollutants, pesticides, organic solvents, and microbes, have long been implicated in PD pathogenesis.^15,16^ These neurotoxicants can be airborne and enter the human body via the nose. The Braak and subsequent dual-hit hypotheses, along with the body-first versus brain-first models of PD, highlight the olfactory route as a possible gateway for environmental toxicants to reach the brain. This pathway allows toxicants to exert direct effects on the olfactory bulb and relevant brain regions, potentially initiating the spread of α-synuclein pathology. Specifically, trichloroethylene (TCE) and bacterial endotoxin LPS have been shown to facilitate PD development through nasal exposure. Experimental studies have demonstrated that TCE can enter the brain via the olfactory nerves, leading to dopaminergic neuronal degeneration in the substantia nigra, accompanied by mitochondrial dysfunction and oxidative stress.^17,18^ Animal studies involving chronic TCE inhalation in rats and mice have shown significant nigrostriatal dopaminergic degeneration, motor and gait impairments, accumulation of phosphorylated α-synuclein in dopaminergic neurons, and microglial activation in the substantia nigra.^19^ Epidemiological studies have also reported that prolonged TCE exposure is associated with increased PD risk, particularly in occupational settings.^20^ These findings underscore the potential role of the olfactory route in the environmental etiology of PD,^21^ supporting the hypothesis that neurotoxicants may initiate neurodegenerative processes.

Intranasal exposure animal models have become increasingly popular in studying environmental neurotoxicants and PD, as they effectively replicate a real-world route of exposure. For example, intranasal administration of MPTP in rodents models the olfactory route of neurotoxicant exposure and disrupts CNS homeostasis.^22^ Recent research demonstrated that unilateral inoculation of α-synuclein preformed fibrils into the olfactory bulb of nonhuman primates leads to severe α-synuclein pathology in the olfactory pathway and limbic system, accompanied by olfactory bulb atrophy and widespread cerebral glucose hypometabolism.^23^ These findings suggest that α-synuclein pathology can propagate from the olfactory bulb and spread PD-like pathology to relevant brain regions, reinforcing the critical role of the nasal route as a gateway for environmental neurotoxicants in PD development.

Our study provides further provocative evidence for this possibility and offers a potential explanation for the asymmetry observed in PD clinical onset and neuronal degeneration. Nasal septum deviation is common in humans and is associated with asymmetry in the olfactory pathway. This study presents the novel and intriguing possibility that nasal septum deviation may increase unilateral exposure to neurotoxicants (illustrated in figure 3), leading to differential CNS exposures and asymmetry in PD pathogenesis. For most patients with nasal septum deviation, we found more pronounced dopaminergic neuronal loss on the dominant side of the deviated septum compared to the contralateral side. However, it is expected that some patients did not exhibit this pattern, as PD has a complex etiology and natural history. It is also plausible that in some cases, PD pathogenesis may originate in the gut or directly in the brain, with minimal involvement of the olfactory pathway. Given the critical role of the olfactory pathway in PD, the development of nose-to-brain drug delivery systems using nano-sized carriers to encapsulate therapeutic agents holds significant promise as a potential intervention for PD.^24,25^ Additionally, the olfactory epithelium, located in the upper part of the nasal cavity, is a mucosal tissue that has recently been identified as a potential pathway for brain-targeted therapeutic delivery through intranasal administration. Research has shown that intranasal administration through the olfactory epithelium provides an efficient route for delivering therapeutics directly to the brain, bypassing the blood-brain barrier and offering a promising approach for brain-targeted treatments.^26^

The etiology of most PD cases likely results from a complex interplay of genetic, environmental, and aging factors.^27,28^ Aging, genetic susceptibility, and environmental influences each play pivotal roles in PD development, with genetic risk factors potentially heightening sensitivity to other factors. Additionally, gene-environment interactions may modulate the effects of neurotoxicants in PD.^17,29^ Heterozygous mutations in PINK1 and Parkin have been linked to PD onset, suggesting they may increase susceptibility to environmental neurotoxicants.^30^ Our analysis of sporadic PD patients revealed a stronger correlation between nasal septum deviation and severe dopamine deficits on the same side, supporting the role of environmental factors in PD lateralization.

This study has several limitations. First, the analyses are cross-sectional and provide no direct evidence of a potential causal link between asymmetric nasal environmental exposure and dopaminergic neuronal loss. Future research should directly test this hypothesis in animal models. Second, this study had limited covariate data and no data on speculated environmental exposures of interest. Nevertheless, we could not identify a major confounding factor underlying both the asymmetries of the nasal cavity and dopaminergic neuron loss. Finally, while the PPMI includes both sporadic and genetic PD cases, we lack genetic data for patients in our local cohorts. However, these three cohorts differ significantly in study settings and patient characteristics, which supports the validity and generalizability of our findings.

In conclusion, this cross-sectional, multi-center observational study proposes a potential mechanism for the asymmetry observed in PD onset and dopaminergic neuronal degeneration, emphasizing the role of the nasal and olfactory pathways in PD pathogenesis. Future research should investigate the nasal pathway as a promising target for PD prevention and treatment.^26^

## Contributors

JianW, CTZ, and MT conceptualised and designed the study. JingW, FTL, JJG, JianW, CTZ, and MT verified the data, and take responsibility for the integrity of the data and the accuracy of their analysis. All authors were involved in data acquisition, analysis, or interpretation. JingW, XNL, YXZ, JFL, CSL, JFL, YYZ, and YCZ contributed to the formal analysis. JingW, FTL, JJG, YMS, WBY, YLT, JJW, HLC, JianW, CTZ, and MT were involved in analysis, or interpretation. JingW, FTL, and JJG drafted the manuscript. JianW, CTZ, MT, and FTL obtained funding. JianW, CTZ, and MT supervised the study. All authors had full access to the data in the study and contributed to revision and editing of the manuscript. All authors contributed to the review and editing of the final version of the manuscript.

## Data sharing

Patient−level data collected for this study will not be made publicly available. The data that support the findings of this study are available from the corresponding author (Jian Wang) upon reasonable request.

## Data Availability

All data produced in the present study are available upon reasonable request to the authors.

## Acknowledgments

We thank the study participants, their families, and the care providers (all staff who had interactions with the study participants, eg, PET imaging technicians, clinicians, nurses, receptionists) who made this study possible. This work used the computational resources of the CFFF platform of Fudan University, and we also acknowledge this platform. We would like to thank Tzu-Chen Yen for the valuable assistance in revising this manuscript. This work received financial support from the Shanghai Municipal Science and Technology Major Project (grants No. 22JC1410402), National Natural Science Foundation of China (grants No. 82171421, 82371432, 92249302, 82272039, 82021002, 81971641, 81902282, 82171252, and 82371266), National Health Commission of China (grants No. Pro20211231084249000238), the Research project of Shanghai Health Commission (grants No. 2020YJZX0111), the Clinical Research Plan of SHDC (grants No. SHDC2020CR1038B), and the STI2030-Major Projects (grants No. 2022ZD0211600). Data used in the preparation of this article were obtained [on June, 03 2024] from the Parkinson’s Progression Markers Initiative (PPMI) database (www.ppmi-info.org/access-data-specimens/download-data), RRID:SCR 006431. PPMI – a public-private partnership – is funded by the Michael J. Fox Foundation for Parkinson’s Research and funding partners, including [list the full names of all of the PPMI funding partners found at the PPMI Website www.ppmi-info.org/about-ppmi/who-we-are/study-sponsors/].

## eMethods

### Detailed neuroimaging methods Imaging acquisition protocol

Cerebral PET/CT dopaminergic imaging was conducted at Gamma Campus of Huashan Hospital using a Biograph 64 scanner (Siemens, Erlangen, Germany) with either ^11^C-CFT or ^18^F-DTBZ as tracers to assess presynaptic dopaminergic dysfunction (DAT or VMAT2). The process began with a CT scan lasting between 18 and 30 seconds, set at 120 kVp and 300 mAs, and featuring a slice thickness of 3·0 to 5·0 mm. Following the intravenous administration of 370 MBq of ^11^C-CFT, a 15-minute PET scan was conducted in a three-dimensional mode at the 60-minute post-injection mark. Alternatively, after administering 370 MBq of ^18^F-DTBZ, a 20-minute scan was performed 90 minutes post-injection. The PET data were subsequently reconstructed using the TrueX algorithm. Subsequently, an all-pass filter was applied to a 336 × 336 matrix for further analysis.

In addition, cerebral PET/CT dopaminergic imaging was conducted at Hongqiao Campus of Huashan Hospital using a uPMR790 PET-MR or uMI780 PET-CT (United Imaging Healthcare, Shanghai, China) with ^18^F-FP CIT as tracers to assess presynaptic dopaminergic dysfunction. The process began with a CT scan lasting between 18 and 30 seconds, set at 120 kVp and 300 mAs, and featuring a slice thickness of 3·0 to 5·0 mm. Following the intravenous administration of 370 MBq of ^18^F-FP CIT, a 20-minute scan was performed 90 minutes post-injection. The PET data were subsequently reconstructed using the TrueX algorithm. Subsequently, an all-pass filter was applied to a 336 × 336 matrix for further analysis.

### PET imaging preprocessing and quantitative analysis

PET images were subjected to spatial normalization, aligning them to the Montreal Neurological Institute’s brain space. Following this, a 3D Gaussian filter with a full width at half maximum of 10 mm was applied for smoothing. The process utilized Statistical Parametric Mapping 12 (Wellcome Trust Centre for Neuroimaging, http://www.fil.ion.ucl.ac.uk/spm/) run on MATLAB R2018a for Windows (The MathWorks Inc; https://www.mathworks.com).

The putamen specific binding ratios (SBRs) was calculated (putamen/occipital-1). We only studied the putamen, since nigrostriatal degeneration affects the putamen before the caudate nucleus, and is therefore a more sensitive measure.^1,2^ Similarly, to previous studies, we obtain an asymmetry index according to the relative difference in SBR.^3,4^ For each bilaterally paired measurement of putaminal dopaminergic deficit, we used right (R) and left (L) hemispheric measurements to calculate the asymmetry index of putamen 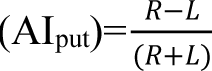. This formula for AI calculation has been well established.^5,6^

In this study, we performed a statistical analysis of the healthy controls, beginning with the calculation of the mean and standard deviation (SD). The healthy controls (n=176, 85 males and 91 females) which were enrolled from HPPI dataset, with an average age of approximately 60 years (58·22±13·75). The SD was utilized as a crucial statistical measure to quantify the dispersion of data points. To better characterize the distribution of the data, we used the SD to define the range of deviations from the mean. Specifically, we calculated the mean of the data and used the standard deviation to determine the distribution range. The graph illustrates the mean, as well as the positions corresponding to the mean±2 SD. This visualization facilitates the observation of data concentration and the identification of potential outliers. eFigure 2A presents the original data distribution, while eFigure 2B depicts the distribution after taking the absolute values of the data. Our calculations yielded a standard deviation of 0·0352, indicating that most of the data points fall within the range of the mean±0·0352. This finding suggests that the data are relatively concentrated around the mean, and the SD offers a precise measure of the variability within the dataset. Based on this analysis, we set the cut-off value as 0·04, ensuring that it captures approximately 95% of the data distribution and helps in identifying potential outliers effectively.

Based on the above analysis, significant dopamine asymmetry in this study was defined as: AI_put_≤−0·04 or AI_put_≥0·04 for primary and internal validation cohorts, and AI_put_≤−0·08 or AI_put_≥0·08 for external validation cohort (as reported by Knudsen et al.^7^).

All SBRs and asymmetry index included in this paper are based on the directly measured SBRs, and are thus not z-scored quantities.

**eFigure 2.**
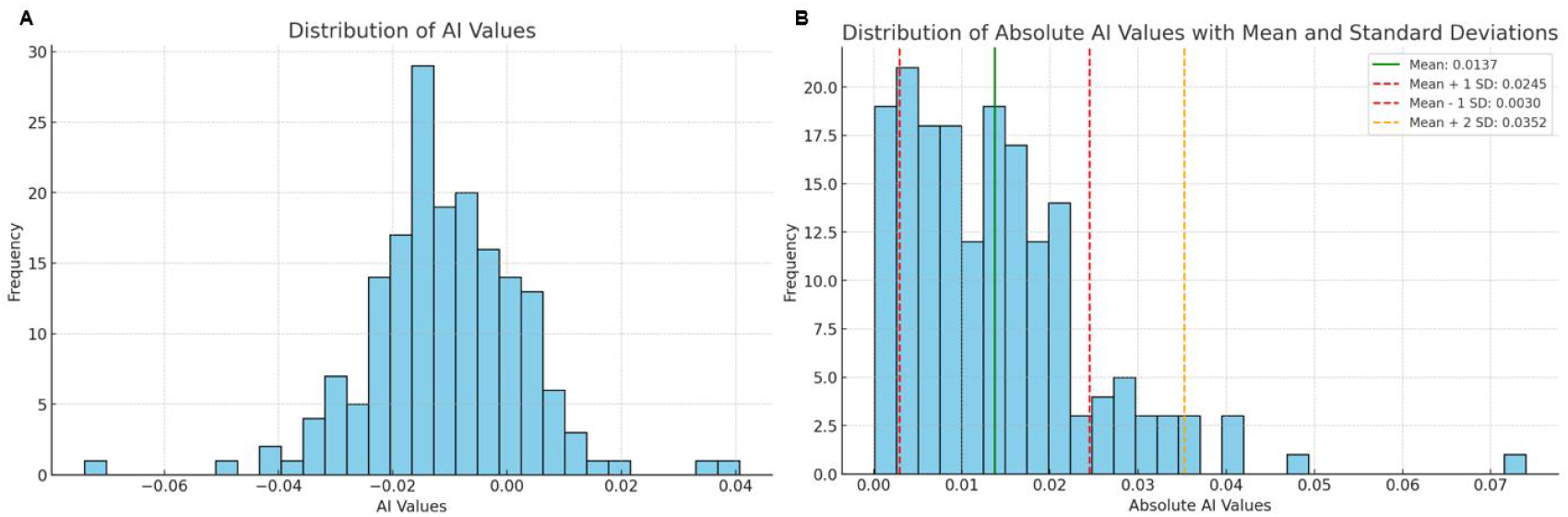
Distribution of AI values. A show the distribution of AI values of all raw data. B presents the distribution of absolute AI values along with the mean and standard deviation lines, which help in understanding the data’s central tendency and variability. AI=asymmetry index.

### Details of CT imaging analysis

#### Visual reads of nasal septum in CT imaging

For the evaluation of the nasal septum, we usually using the following steps: Firstly, reference lines on axial (eFigure 3A) and sagittal position (eFigure 3C) MPR images are used to correct the measurement plane. Secondly, the septal deviation was evaluated the direction of deviated nasal septum in coronal (eFigure 3B) sections MPR images at the point of maximal septal deviation by two expert readers, who were blind to the patients’ clinical and dopamine deficit features.^8^

**eFigure 3.**
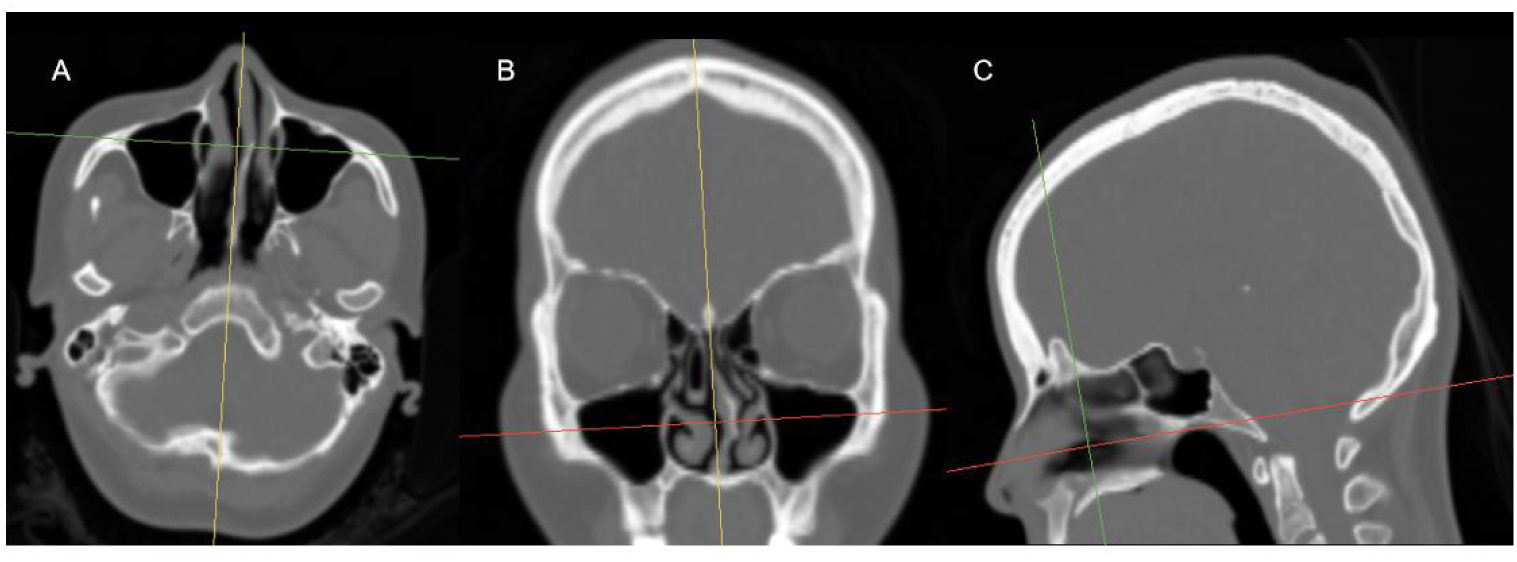
Deviated nasal septum visual assessment. A−C was one patient’s assessment as unilateral deviation (deviating to either the left side, i.e., concave side of nasal septum deviation on right, dominant side of the nasal cavity on right).

To evaluate the inter-rater reliability between two raters, we employed Cohen’s kappa coefficient (κ) and percentage agreement. Cohen’s kappa is a statistical measure used to assess the degree of agreement between two assessors while accounting for the agreement that may occur purely by chance. We also calculated the 95% confidence interval for the κ estimate to provide additional context on the reliability of the calculated agreement level. Cohen κ values are generally interpreted as follows: ①< 0·20: Poor agreement; ②0·20 − 0·39: Fair agreement; ③0·40 − 0·59: Moderate agreement; ④0·60 − 0·79: Substantial agreement; ⑤0·80 − 1·00: Near perfect agreement.

The Cohen’s κ of external validation cohort was 0·96 (95%CI, 0·931 − 0·989), indicating a near perfect agreement between the two raters. The percentage agreement was found to be 97·15%, suggesting that the vast majority of ratings were consistent between the raters. Additionally, a confusion matrix was generated to provide a detailed overview of the classification performance between the two raters (eFigure 4). The matrix demonstrated that most of the ratings were correctly aligned across the categories, with only a small number of discrepancies. These results indicate a high degree of consistency and reliability in the ratings provided by both evaluators.

**eFigure 4.**
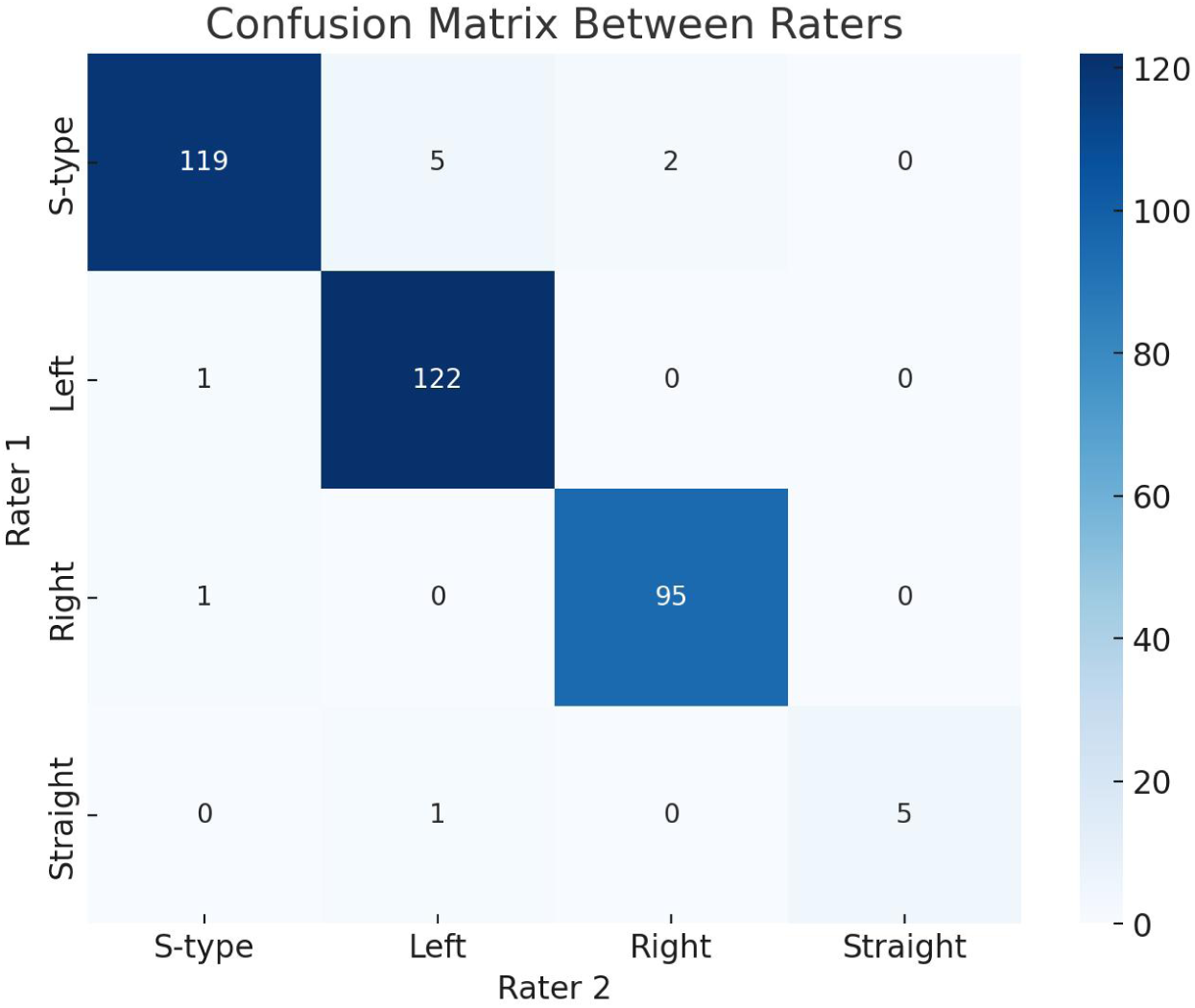
Confusion matrix between two raters. This figure illustrates the agreement between two raters across different categories. The heatmap shows the frequency of ratings for each combination of categories. Darker blue indicates higher agreement, while lighter colors indicate fewer occurrences. The diagonal cells represent instances where both raters provided the same rating, highlighting strong agreement. The off-diagonal cells indicate disagreement between the raters, with the counts specifying how often such mismatches occurred.

### Nasal cavity segmentation and phenotype extraction

#### Manual segmentation

All the CT scans are annotated with two anatomical structures including the left nasal cavity, right nasal cavity. The annotation process involved pixel-wise annotation, where each pixel was carefully labelled to identify to different structures or background. The manual annotation and refined procedure of segmentation is adopted in 3D Slicer 5.6.2 (Asan Medical Center Nuclear Medicine Image Quantification Toolkit of Excellence), a software tool available at https://www.slicer.org/. Air thresholding was set at −400 to 1000 Hounsfield units (HU) for all scans. Then, the filter module (i.e., LaplacianSharpeningImageFilter) was used to highlight regions with rapid intensity changes and thus enhance the edges, followed by the Segment Editor module for segmentation Due to the high annotation cost of annotating large-scale 3D medical image segmentation datasets, we adopt a human-in-the-loop procedure to facilitate the annotation procedure.

Step 1. Manually annotate a small subset (eFigure 5). To begin, we manually annotate 21 cases from total data from scratch for the development of the initial segmentation model. Three highly trained junior annotators perform the annotation.

Step 2. Develop the initial version of the segmentation model. Based on the annotated subset, we train a segmentation model based on nnUNet^9^ as the initial segmentation model. Then the initial segmentation model is utilized to infer the remaining cases and generate pseudo-segmentation masks.

Step 3. Refine the coarse segmentation masks. Building upon the output of the initial segmentation model, the coarse segmentation masks of remaining cases are then manually refined by the annotators.

Step 4. Re-train the model for cross-validation. After acquiring the annotation, we split the data into five folds for cross-validation.

Step 5. Checking and correcting by senior experts. To ensure the accuracy of annotation, after cross validation, cases with lower scores are double-checked by the senior experts.

**eFigure 5.**
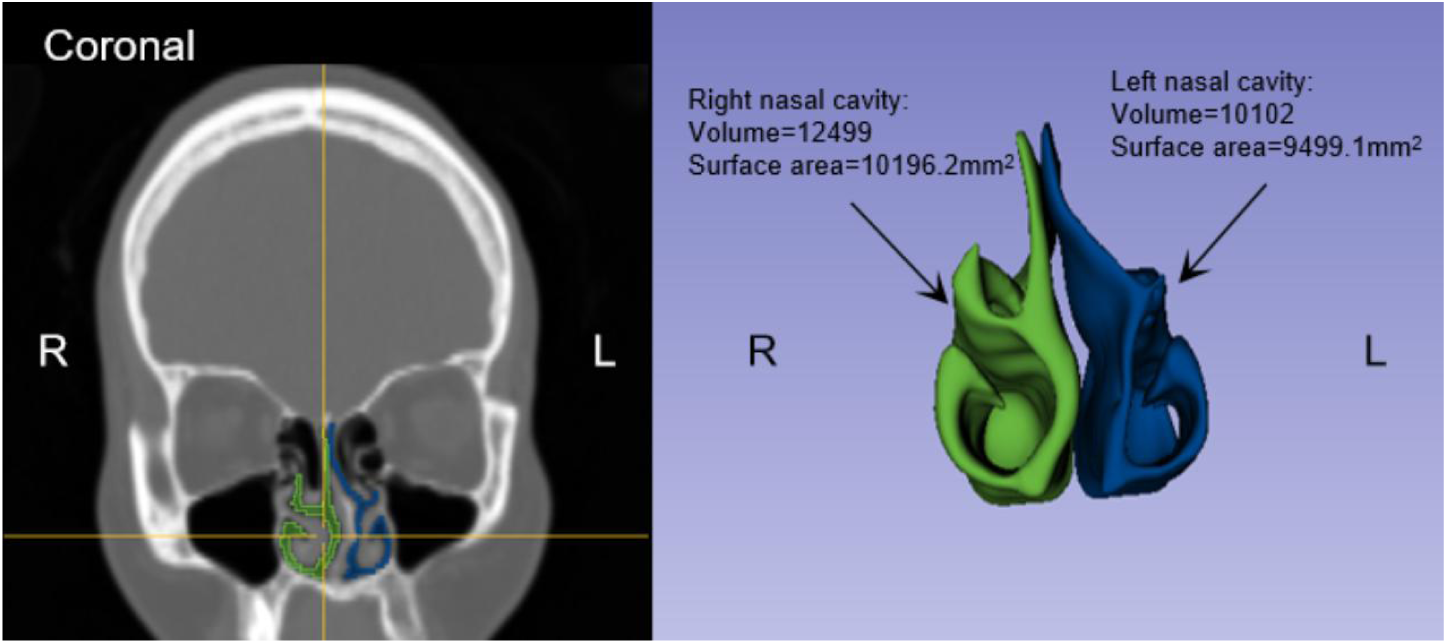
Segmentation of nasal cavity. This is the nasal cavity segmentation image of Patient B in the main text.

### Interatively segment and refine labels

#### Network Architecture

The network architecture is building based on U-Net characterized by its encoder-decoder structure with skip connections, which facilitates the restoration of spatial information lost during the encoding process. The base number of features for the U-Net is set to 32, with the maximum number of features in the deepest layer reaching 320. The network is designed with a series of convolutional layers, each followed by a ReLU activation function, except for the last layer which utilizes a softmax activation for segmentation purposes. The convolutional layers are composed of varying kernel sizes, predominantly 3*3*3, to capture anisotropic features within the medical images.

#### Preprocessing

Prior to training, the dataset undergoes a series of preprocessing operations. The images are normalized using a CT Normalization scheme without the application of a mask for normalization. The median image size in voxels is maintained at 148*512*512 with a spacing of 1·5*0·5859375*0·5859375. The dataset is also subjected to augmentation techniques to enhance the robustness of the model. These include random rotations, scaling, and the introduction of Gaussian noise and blur. A multiplicative brightness transform and contrast augmentation are applied to simulate variations in image acquisition. Additionally, a low-resolution simulation is performed to account for the variability in image quality.

#### Training Protocol

The training is conducted on a NVIDIA A100-SXM4-80GB GPU with the use of mixed-precision training through the GradScaler. The training dataset is loaded in batches of size 2, with a total of 250 iterations per epoch. The validation dataset follows a similar loading pattern but with 50 iterations per epoch. Data augmentation during training includes spatial transformations, such as rotations and scaling, with a probability of 0·2 and 0·2 respectively. Gaussian noise is introduced to the data with a variance ranging from 0 to 0·1, and Gaussian blur is applied with a sigma range of 0·5 to 1·0.

#### Hyperparameters

The initial learning rate is set to 0·01, and the optimization is performed using the Stochastic Gradient Descent (SGD) optimizer with a momentum of 0·99 and weight decay of 3e−05. The learning rate scheduler employed is a polynomial decay (PolyLRScheduler), which adjusts the learning rate across epochs to prevent overfitting and to enhance convergence. The loss function is a composite of the Cross-Entropy (CE) loss and the Memory-Efficient Soft Dice (DC) loss, encapsulated within a Deep Supervision Wrapper to ensure consistent performance metrics and to facilitate multi-level loss computation.

#### Visual assessment the quality of segment

The segmented data is checked visually and, in the event of a segmentation error, a manual adjustment is made.

#### Assessments of nasal cavity phenotypes

The deviation phenotypes, including the volume and surface area of both sides of the nasal cavity, were extracted using 3D Slicer software (Radiomics; Asan Medical Center Nuclear Medicine Image Quantification Toolkit of Excellence).

#### Nasal computational fluid dynamics simulation

The patterns of nasal computational fluid dynamics (CFD) in a PD patient (Patient B) with a deviated nasal septum, showing the dominant side of the nasal cavity on the right.

To gain more quantitative insights into this problem, a CFD simulation was conducted. CT scan data were used to extract and reconstruct the airway boundaries, from which an unstructured tetrahedral mesh of the nasal cavity was generated. This mesh contains approximately 190,000 cells for both the left and right airways, with a maximum edge length of 0·6 mm. To accurately model airflow behavior near the nasal cavity walls, three layers of boundary cells were added. An overview of the mesh is provided in eFigure 6.

The CFD solver employed in this study is a steady laminar flow solver. Air density is set at 1·18415 kg/m³, with a dynamic viscosity of 1·85508 × 10⁻⁵ Pa.s. The inlet pressure is specified as zero to serve as a reference, while the outlet pressure is set at -500 Pa to simulate the inhalation process.

Solid particles have a diameter of 2·5 µm and a density of 1200 kg/m³. Particle motion is simulated using a Lagrangian approach, with a one-way coupling for solid-fluid interactions, given that the particles are very small and sparse. The forces on the particles are calculated using the Schiller-Naumann aerodynamic drag model.

The CFD simulation is first performed, achieving a stable and convergent flow field after 3000 iterations. Particle tracking then follows, with particles released at the inlet to trace their paths. Particles freeze upon contact with the wall or upon reaching the outlet. All meshing and simulations were conducted in Star-CCM+ 12.06.011, and the rendering of simulation results was completed in Blender with custom modifications.

**eFigure 6.**
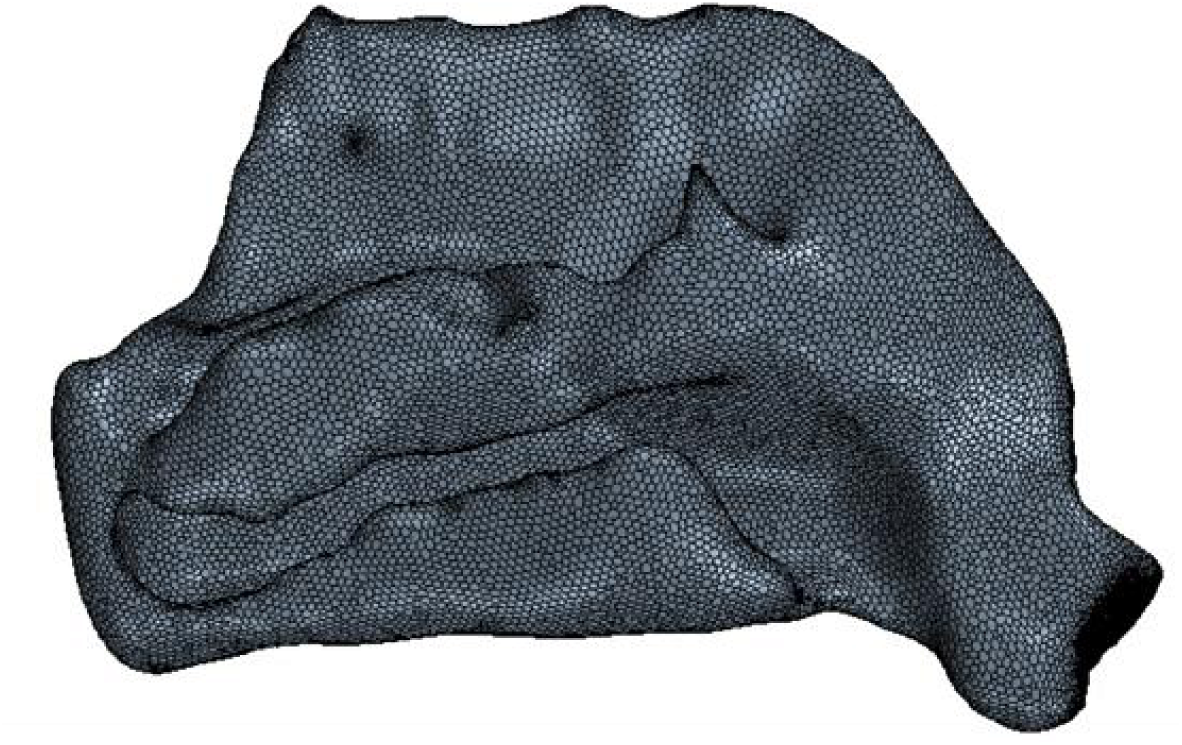
Mesh representation of the domain used for CFD simulations

### Other statistical analysis

#### Phi Coefficient

The Phi Coefficient (φ) is a measure of association between two binary variables.^10^ It means the independent variable and dependent variables are interchangeable. The interpretation for the φ is similar to the Pearson Correlation Coefficient. The range is from −1 to 1, where: 0 is no relationship. 1 is a perfect positive relationship: most of your data falls along the diagonal cells. −1 is a perfect negative relationship: most of your data is not on the diagonal. The interpretation for the φ as followed:

| $\phi$ value | Interpretation |
| --- | --- |
| +0.70 or higher | Very strong positive relationship |
| +0.40 to +0.69 | Strong positive relationship |
| +0.30 to +0.39 | Moderate positive relationship |
| +0.20 to +0.29 | Moderate positive relationship |
| +0.01 to +0.19 | No or negligible relationship |
| 0 | No relationship |
| -0.01 to -0.19 | No or negligible relationship |
| -0.20 to -0.29 | weak negative relationship |
| -0.30 to -0.39 | Moderate negative relationship |
| -0.40 to -0.69 | Strong negative relationship |
| -0.70 or higher | Very strong negative relationship |

## eResults

### Participants

**eFigure 7.**
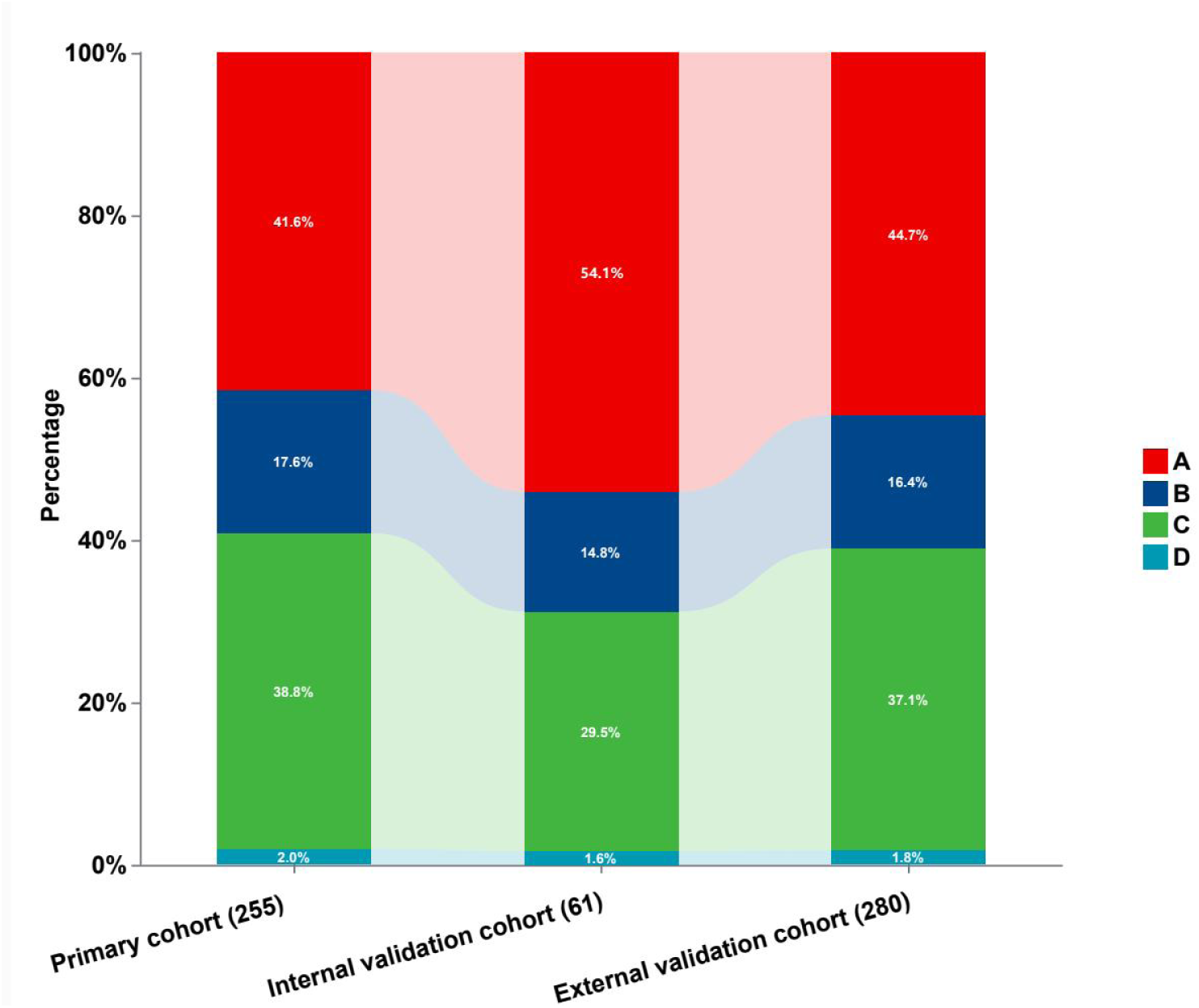
Distribution of different nasal septum patterns in Parkinson’s disease patients with pronounced striatal dopaminergic asymmetry across the three cohorts. (A) The side with a more pronounced striatal dopaminergic deficit was consistent with the dominant side of the nasal cavity. (B) The side with a more pronounced striatal dopaminergic deficit was inconsistent with the dominant side of the nasal cavity. (C) S-shaped deviation. (D) Straight septum.

**eTable 1.** Correlation between the Dominant Side of Nasal Cavity and the Striatal Side of More Pronounced Dopaminergic Deficit based on Hoehn-Yahr Stage

| Dominant side of nasal cavity | Striatal side of more pronounced dopaminergic deficit | | | *P* value ($\phi$) |
| --- | --- | --- | --- | --- |
|  | left | right | Total |  |
| Early-stage patients (Hoehn-Yahr stage=1) |  |  |  | <0·001 (0·48) |
| Left | 22 (36%) | 8 (13%) | 30 |  |
| Right | 8 (13%) | 23 (38%) | 31 |  |
| Total | 30 | 31 | 61 |  |
| Lately-stage patients (Hoehn-Yahr stage=2–5) |  |  |  | <0·01 (0·35) |
| Left | 25 (28%) | 15 (17%) | 40 |  |
| Right | 14 (16%) | 36 (40%) | 50 |  |
| Total | 39 | 51 | 90 |  |

**eFigure 8.**
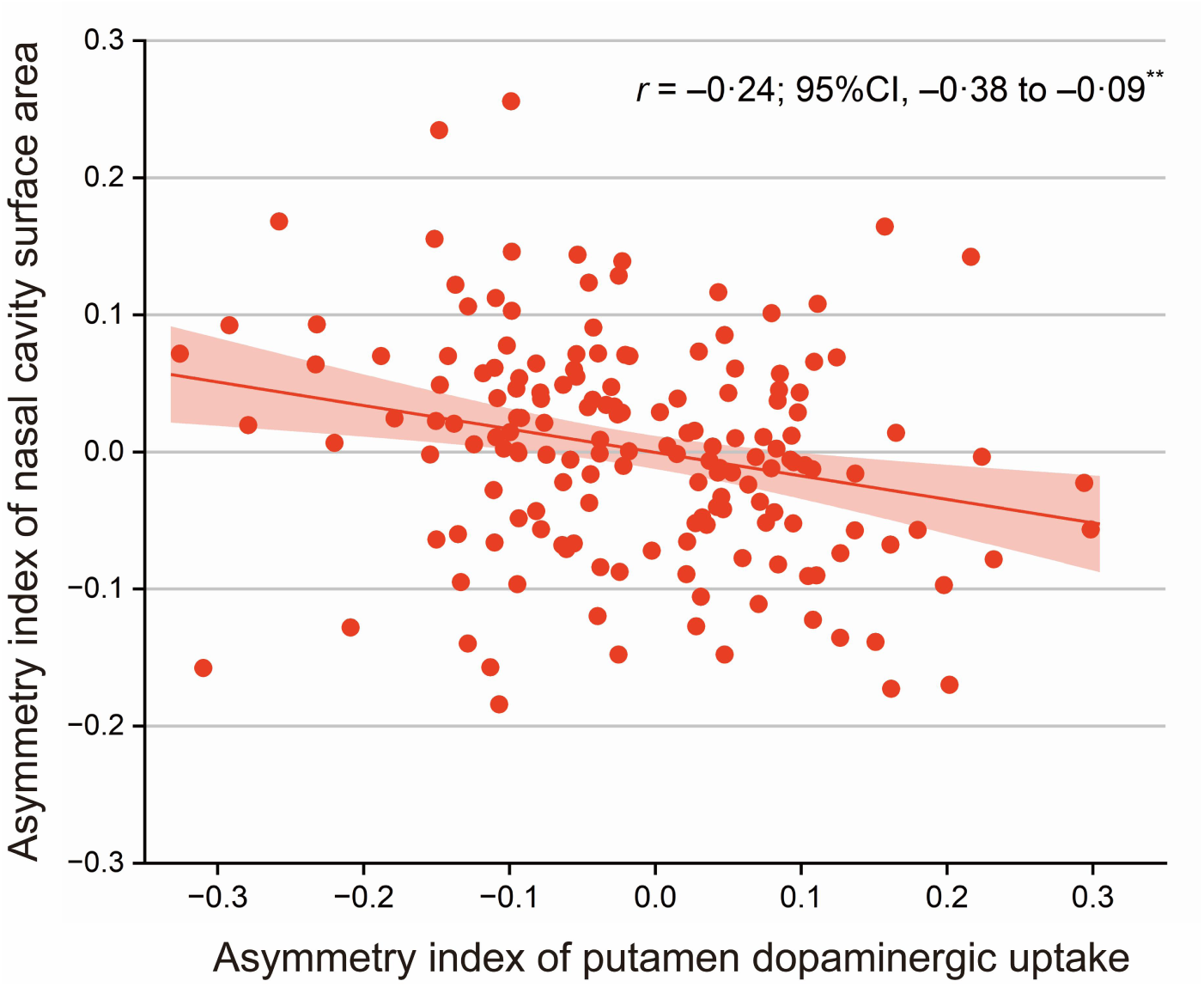
Association between the Laterality Index of Nasal Cavity Volume and Laterality Index of Putamen Dopaminergic Uptake in the Primary Cohort ** p<0·01

